# A comparison of two open-source methods for molecular phylogenetic analysis of Human Immunodeficiency Virus (HIV): Hypothesis testing using Phylogenetics (HyPhy) and Molecular Evolutionary Genetics Analysis (MEGA)

**DOI:** 10.1101/2025.09.28.25336847

**Authors:** Craig A Davis, Simon Reid, Charles F Gilks

## Abstract

We compared two widely used methods for Human Immunodeficiency Virus (HIV) molecular phylogenetic analysis (MPA) - Hypothesis testing using Phylogenetics (HyPhy) and Molecular Evolutionary Genetics Analysis (MEGA) – to consider which could strengthen surveillance and better target prevention interventions in Queensland, Australia.

HIV *pol* sequences generated for drug resistance testing were linked to de-identified case reports in the state-wide register of notified HIV cases. The study used MEGA, 6.0 patristic distance ≤1.5% and HyPhy, patristic distance ≤2% to identify molecular transmission clusters. We compared time taken to perform each analysis; the effectiveness of identifying sequences that clustered; the efficiency of identifying clusters and their sizes; and the ability simply to illustrate cluster networks and their evolution over time.

Of 1776 unique sequences identified 1,563 (88.5%) were linked to a notification record. Analysis with HyPhy (30 minutes) was 600 times faster than MEGA (324 hours). With HyPhy, 1084 (61.4%) sequences clustered compared to MEGA where 595 (33.7%) clustered making HyPhy 54% (595/1084) more effective. Overall, HyPhy identified 82 more transmission clusters than MEGA (266 versus 184) performing 45% (82/184) more efficiently. In size terms, HyPhy found 565 sequences clustered in 50 moderate or large clusters with MEGA finding 261 sequences in 21 moderate and 2 large clusters: of these clusters, 43 were identified by both methods but with MEGA nearly half (20; 47%) were small. HyPhy also more efficiently established cluster size. The HyPhy network cluster maps can more simply illustrate molecular transmission clusters, include patient characteristics, show their timelines and are easier to update than the cluttered MEGA circular phylogenetic trees.

We are confident that HyPhy is better suited than MEGA to use in the Queensland context to generate comprehensive network transmission cluster maps for monitoring HIV transmission and to translate near real-time phylogeny data into action for targeted prevention interventions.

## Introduction

With the widespread availability and increasing simplicity of sequencing technology, pathogen isolates are now regularly sequenced for clinical and epidemiological purposes. Molecular phylogenetic analysis (MPA) describes similarities and differences in sequences between clades, groups of organisms that share a common ancestor. A phylogenetic tree to represent the evolutionary relationship is created by linking the least distant pairs of sequences followed by successively more distant sequences. Cladistic branching tree-like diagrams are produced where the leaves are individual sequences and the patristic distance, the length of the branches from tip to next tip, is a measure of the divergence between individual sequences. Four distinct steps are involved in MPA: acquiring the sequence data; aligning the sequences; estimating a tree from the aligned sequences; and presenting the phylogenetic relationship in a clear, understandable visual representation (1).

In the case of human immunodeficiency virus (HIV), in high-income countries like Australia clinical isolates are routinely sequenced to identify antiretroviral drug resistance to guide treatment; this involves the relatively conserved *pol* region of the HIV genome that encodes protease and reverse transcriptase, the primary targets of most antiretroviral medicines. The *pol* sequences that are generated can be used for MPA which can be particularly informative when sequence data are linked to risks factors in notification reports and with basic clinical data. This has enhanced our ability to characterise transmission networks, identify transmission clusters and describe cluster dynamics over time (2). Surveillance systems can be applied to generate near real-time data for identifying potential historic and ongoing HIV transmission clusters and thus relatively easily translate phylogeny into public health action for HIV control (3, 4).

There are several different open-source programmes that can easily be downloaded onto desktop computers and used in a standard operating environment. These include MEGA (Molecular Evolutionary Genetic Analysis), HyPhy (HYpothesis testing using PHYlogenies), BEAST (Bayesian Evolutionary Analysis by Sampling Trees), HIV-TRACE (Transmission Cluster Engine), PhyML (Maximum-Likelihood Phylogenies), Cluster Picker and PHYLIP (PHYLogeny Inference Package) (1, 5–10). These use different methods to reconstruct HIV phylogenies: the hierarchical neighbour joining (NJ), maximal likelihood (ML) across all possible tree reconstructions and Bayesian, assuming a constant rate of evolution across branches.

The maximum likelihood method, which infers a tree based on the minimum number of changes required to explain the data, is widely used because it adds branches at different levels on the phylogenetic tree in a stepwise fashion with each level evaluated for best-fit before the next branch. It is however quite time-intensive compared to other methods (11). The robustness of a tree is improved by bootstrapping (resampling) techniques. A bootstrap value of ≥98% and an intra-cluster genetic patristic distance, a measure of similarity to the nearest neighbour, of ≤1.5% or ≤2.0 is generally considered to represent true linkage between sequences (12).

HIV type 1 (HIV-1) is the predominant type globally; it has various subtypes. Several molecular epidemiology studies have used MEGA to define HIV-1 subtype diversity patterns and for phylogenetic analysis in order to construct possible HIV transmission networks, including from Australia (13), Canada (14), Mexico (15) and South Africa (16). Other studies have used HyPhy for molecular transmission network analysis (17–19) and to identify HIV transmission clusters in near real-time (3, 20, 21). Some groups have used HIV-TRACE to derive HIV transmission networks and clusters, using HIV *pol* and *gag* sequences (22, 23); one study sequenced *gp41* and used HIV-TRACE with Cluster Picker (24). The PhyML method has been used by one group in Brazil with both *env* and *pol* sequences (25).

In Australia, HIV infection (both with HIV-1 and HIV type 2) has been notifiable since 1984; almost all notifications have been with HIV-1. The data collected form part of the national HIV minimum data set for Australia. Since 2008, an HIV-1 pol RT-PCR assay amplifying a partial *pol* HXB2 gene sequence has been used as part of routine clinical management for monitoring HIV-1 drug resistance in all newly notified cases and on patients already in care with suspected drug resistance. *Pol* sequence data are archived by Queensland Health Pathology Services.

As part of a state-wide study of the molecular epidemiology and phylogeny of HIV, two well-established open-source methods for MPA, HyPhy and MEGA, were compared using readily available HIV-1 *pol* sequences to consider which could be used in the Queensland context to generate near or real-time surveillance data for monitoring HIV transmission to translate into action for more targeted HIV prevention interventions.

## Methods

### Data Sources

All notification data and genotype resistance testing (GRT) sequences were obtained from the Queensland Department of Health (QDoH). Deidentified HXB2 sequences generated for genotypic HIV-1 drug resistance testing carried out by QDoH over a six year period from 1^st^ January 2008 to 31^st^ December 2013 were linked to de-identified HIV case reports in the state-wide register of notified HIV cases. Deidentified HIV notifications recorded in the QDoH Notifiable Conditions System (NOCS) for the period 1^st^ January 1984 to 31^st^ December 2013 were included in the study.

### GRT Sequences

Laboratory records were obtained from the Queensland HIV Reference Laboratory (QHIVRL), part of the Queensland Health Pathology Services (QHPS). Plasma specimens underwent HIV-1 subtyping and genotypic resistance testing (GRT) using the ViroSeq® HIV-1 Genotyping System assay using Sanger real-time polymerase chain reaction (RT-PCR). GRT targeted the partial *pol* gene including full coverage of the protease enzyme (codons 1-99) and two-thirds coverage of the RT enzyme (codons 1-335). Each test produced a nucleotide sequence in the form of a text-based formatted file (FASTA universal standard). Laboratory identifiers (test date, FASTA file name, name-code, birth date, laboratory number and specimen collection date) were used to identify baseline from duplicate sequences; only the initial baseline sequence was included. The laboratory identifiers were used to improve identification of notification and clinical records for linkage and to aggregate the testing history of each case. The time to GRT was measured as the difference between the date of the earliest HIV diagnosis and the date of the first baseline GRT.

### Notification Data

Notification data items included: date of first HIV and Acquired Immunodeficiency syndrome (AIDS) diagnoses; age at HIV and AIDS diagnoses; age on 31^st^ December 2013; if relevant deceased date and age at death; sex (self-identified); indigenous status; country and region of birth; local area statistical division; HIV risk exposure; HIV seroconversion status; and late HIV diagnosis. HIV risk exposure is based on patient -reporting of exposure to the diagnosing clinician and is recorded as up to two data items with priority given to either or both the type of sexual contact and injecting drug use (IDU).

Also included in the notification data were HIV viral load and CD4 counts measured over the 2001 to 2013 period. Viral load was recorded for analysis as <400 or 400+ copies/ml (copies of HIV RNA per millilitre of blood). The CD4 count was recorded as <200 or 200 or more cells/mm^3^ (cells per cubic millimetre of blood). In our setting, individuals with a CD4 count <200 cells/mm^3^ are defined as having immunological AIDS; the normal adult CD4 count reference range is between 500 and 1,600 cells/mm^3^.

### Data Linkage

Linkage between notification and GRT records was carried out by the Communicable Diseases Branch (CDB), QDoH. Notifications were used as the primary reference; name-code, birth date, and sex were used as personal identifiers in common between the data sets. Clerical review was used to inspect uncertain matches. Sequence (FASTA-formatted) files were provided directly by CDB. All other linkage was carried out by the Statistical Analysis and Linkage Team (SALT) using notifications as the primary dataset. Due to the limited personal identifiers, only deterministic linkage methods were used to connect notifications, prescriptions, and laboratory (excluding sequence) records. No personal identifiers were provided for analysis.

### Molecular Phylogenetic Analysis

The two chosen open-source phylogenetic analysis methods were compared using standard personal computer hardware and software operating environments. MEGA was operated in a Windows 7 (and version 10) operating environment and was carried out on a Toshiba Kira Intel® Core™ i7-4500U 2.4GHz, 8GB RAM, Intel HD Graphics. HyPhy was operated in an OS X 10.10 Yosemite (later upgraded to OS X 10.11 El Capitan) environment and was carried out on a MacBook Pro Intel Core i5 2.7 GHz, 8Gb RAM, Intel Iris Graphics 6100 1536 MB.

The study used MEGA version 6.0 (26), and followed the method described by von Wyl and colleagues at the standard patristic distance of ≤1.5% (27). An unrooted neighbour-joining maximum-likelihood tree using a general time reversible (GTR) model gamma distributed with invariant sites, bootstrap resampling with 1000 replicates, nearest-neighbour-interchange (NNI) and maximum parsimony was constructed. Seventy-one reference sequences, including 15 subtype (A-K) reference sequences and 56 CRF reference sequences, were included in the MPA analysis to facilitate identification of similar sequences in the dataset. BioEdit ClustalW alignment was carried out prior to MPA on all sequences (28).

Sequences were classified as clustered if the shortest patristic distance to another sequence was below the cut-off in 80% or more of the bootstrap trees. Those which met the criteria for clustering were displayed in the results; non-clustering sequences were excluded. Clusters were identified manually from the printed output calculated as the (additive) distance between sequences; manual calculation is required where additive patristic distances for immediately adjoining branches was less than the cluster criterion. Two or more sequences clustering around a common proximal node with ≥99% bootstrap support and at least 1 neighbour with patristic ≤1.5% substitutions per branch length were named. Clusters were categorised as small (2 to 4 sequences), moderate (5 to 14 sequences), or large (15+ sequences). Clusters were also described according to duration (years from diagnosis of the first to the most recent case) and growth (average annual increase in cluster size).

HyPhy was developed in 2000 (29) and since then has regularly been updated (8). The study followed the approaches outlined by Poon and colleagues (3, 20). Sequences were translated into amino acids and aligned pairwise against a single HXB2 subtype B reference sequence. Sequences were classified as clustered if their shortest patristic distance (divergence) to another sequence was below cut-off in 80% or more of the bootstrap trees. To control for uncertainty in phylogenetic reconstruction, the method generated 100 bootstrap samples by resampling columns from the alignment at random with replacement. Patristic distances were then extracted from 100 replicate trees reconstructed from bootstrap alignments using FastTree2 (30). Clusters were identified visually from the output and uniquely named and described as above for MEGA.

### Comparison of MPA Methods

Several objective comparisons were made between the two phylogenetic analysis methods including the time taken to perform each analysis; the effectiveness of identifying the number of sequences that clustered; and the efficiency for defining the number and size of the molecular transmission clusters identified. For MEGA a ≤1.5% patristic distance was used; for HyPhy a ≤2.0% patristic distance was used as reference and other patristic distances were explored. The distance between nodes represents similarity of sequences: shorter lines indicating greater sequence similarity. Varying (bracketing) the patristic distance impacts cluster size: smaller patristic distances reduce the number of clustering sequences. Subjective comparisons included the clarity that phylogenetic relationships were visually represented as either circular phylogenetic trees or as molecular transmission clusters; how clearly the graphical representations were able to show sequence relatedness and identity; the ability to describe cluster dynamic and timelines; and the ease with which new sequences can be related to existing molecular clusters for surveillance purposes.

### Statistical Analysis

For statistical comparisons, a range of univariate, bivariate, (including frequencies, cross-tabulations, analysis of trends in distribution and proportions/rates over time, chi-square, Mann-Kendall, Barnard’s) and multivariate (linear and logistic regression, factor analysis) statistical techniques were used as appropriate to identify useful surveillance indicators of HIV care. Analyses were performed using IBM SPSS Statistics for Windows, Version 21.0.

### Approvals

Ethical approval for the study was provided by the Human Research Ethics Committee, Gold Coast Hospital and Health Service (HREC/13/QGC/141) and by the University of Queensland Medical Ethics Research Committee (#2014000047). The data were retrospectively accessed in April and May 2014 for research purposes; all data accessed were deidentified and the study team had no information that could identify individual participants either during or after data collection. The data were provided with a Waiver of Consent that was granted by the UQ Medical Ethics Research Committee. Approval for data access was granted in accordance with Section 284 of the Queensland Public Health Act (2005) (RD005115).

## Results

In Queensland, over the period 1984 to 2013 a total of 6,441 people were notified with HIV and 1758 AIDS diagnoses were made. Most reported cases of HIV were male (90.5%). There were 4,702 (73.0%) notified cases of HIV with a CD4 count recorded at diagnosis: one in five CD4 counts were <200 cells/mm^3^. There were 3,107 (51.8%) notified cases of HIV with a viral load recorded at diagnosis: one in five were <400 copies/ml.

The majority of people reported with HIV infection in Queensland (4636/6441; 72.0%) were born in Australia. The remainder (28.0%) were born elsewhere: 518 (8.0%) were born in Europe, 395 (6.1%) were born in Oceania, 279 (4.3%) were born in Sub-Saharan Africa (SSA), 217 (3.4%) were born in South-East Asia (SEA) and 194 (3.0%) were born in the Americas. More than half of the notified cases (53%) resided in Brisbane, the state capital, at time of diagnosis.

The largest reported risk group for HIV exposure was men who have sex with men (MSM; 75.8%). This group included MSM with no other risk factors (58.7%), bisexual men with no other risk factor (8.7%), MSM who also injected drugs (6.6%), and bisexual men who also injected drugs (1.8%). Heterosexual exposure (8.7% overall) steadily increased over the study period from 1% in 1984 to 19% in 2013. Overall, 4.5% of notifications had no identified exposure risk.

There were 2,243 complete GRT sequence records generated in the study period of which 1,766 (76.1%) were first sequences from individual cases and were available for analysis. Of these, 88.5% (1,563/1766) were linked to a notification record and 203 (11.5%) were without linkage to any notification. Slightly more than half (856; 54.8%) of the linked sequences were generated within one year of first HIV diagnosis, with the majority within 3 months of diagnosis; these were considered baseline. The remainder (707; 45.2%) of the linked sequences were obtained from individuals who had been diagnosed before 2008 and these were considered first sequences. There were 1171 (75%) baseline/first linked sequences from individuals who identified as MSM and 61 (3.9%) from individuals identifying as Indigenous Australians.

Some study sequences were found just once (singleton); depending on the method used, many were found more than once and could be regarded as clustered. Cluster size was categorised as small (2 to 4 sequences), moderate (5 to 14 sequences), or large (15 or more sequences). HIV-1 subtype B predominated (1,393/1766; 78.9%). Of the 373 (21.2%) non-B subtypes, subtype C (166; 9.4%) and CRF 01_AE (116; 6.6%) were the most common. The proportion of subtype B sequences significantly decreased from 85.5% in 2008 to 72.9% in 2013 (p < 0.05), both for sequences linked and without linkage to notification records. There were significantly more subtype B sequences from Indigenous Australians compared with non-B subtype sequences (55 versus 6. OR = 2.38; 95% CI: 1.02 - 5.58).

### MEGA Analysis

The total duration of the analysis of the 1766 study sequences using MEGA with ≤1.5% patristic distance was 13.5 days (324 hours) with alignment taking 38 hours and molecular phylogenetic analysis taking 286 hours. Branch lengths were assessed manually to determine molecular transmission clusters. Cluster labelling was also done manually after analysis. Figure 1 shows a circular unrooted phylogenetic MEGA tree, with clustered subtype B and non-B (D, B/C, C, 01-AE, and others) sequences separately highlighted.

**Figure 1:**
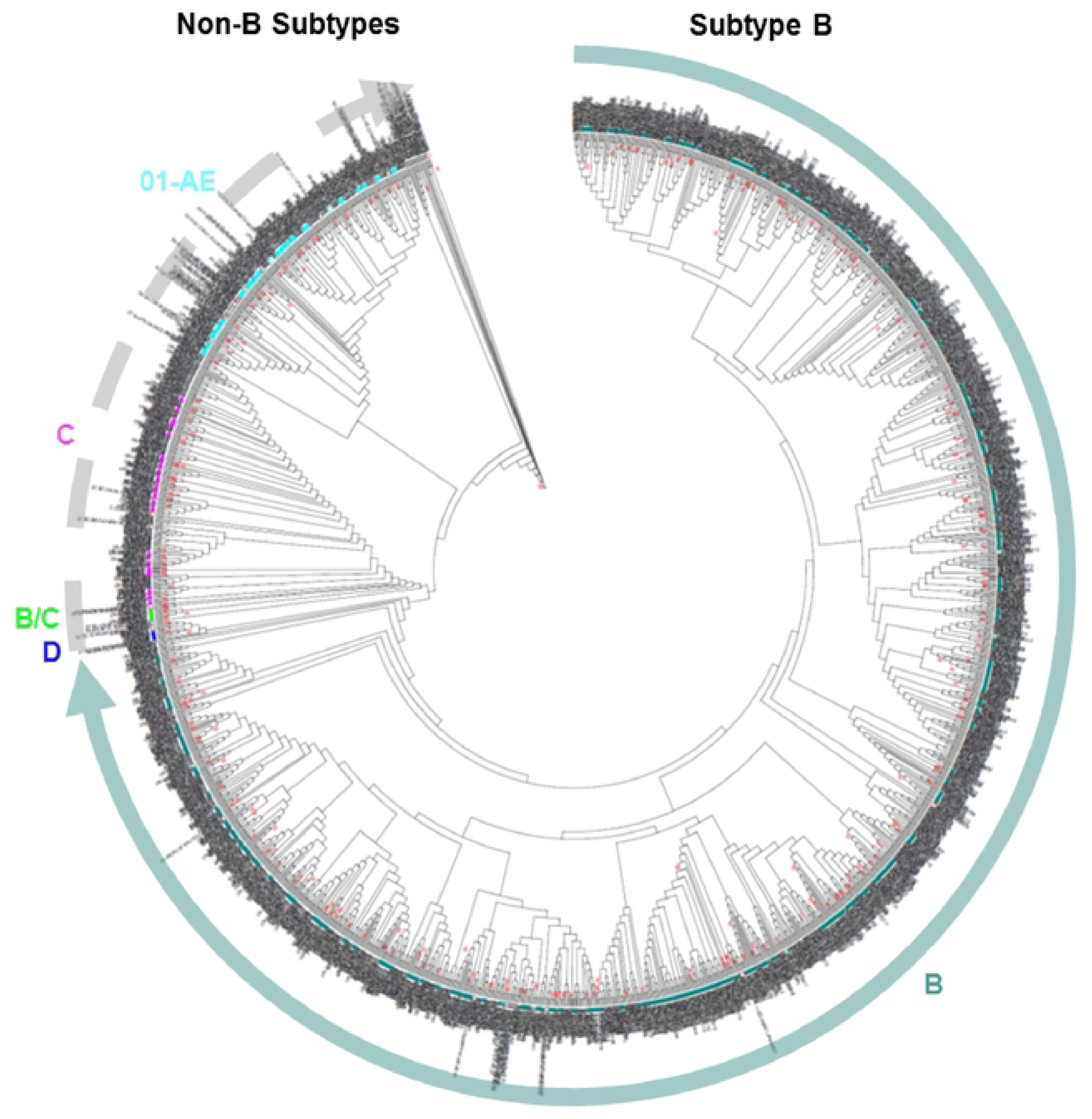
MEGA circular phylogenetic tree including all 1,766 GRT sequences and 72 reference sequences indicating portions which are subtype B and non-8 subtypes. Each sequence is individually labelled (longer labels indicate reference sequences). Branch lengths were used to manually determine molecular transmission clusters (MTCs).

The number and size of the singletons and clusters MEGA identified is depicted in table 1 by subtype, sequence linkage and frequency by HIV-1 subtype. Two-thirds (1,171/1766; 66.3%) of the study sequences were singletons that did not cluster with any other sequence. One-third (595/1766; 33.7%) formed 184 clusters of which 161 were small, 21 moderate and two large. There were 136 subtype B clusters including 82 pairs and 19 moderate or large clusters of five or more sequences, including a cluster with 17 sequences and a larger cluster of 62 sequences. Of the 52 non-B subtype clusters, 37 were pairs and four were moderate; no large clusters were found. There was no significant increased likelihood of clustering (two or more) with subtype B sequences compared with non-B sequences (34.2% versus 31.7%. OR = 1.1; 95%CI = 0.9 - 1.3).

**Table 1:**
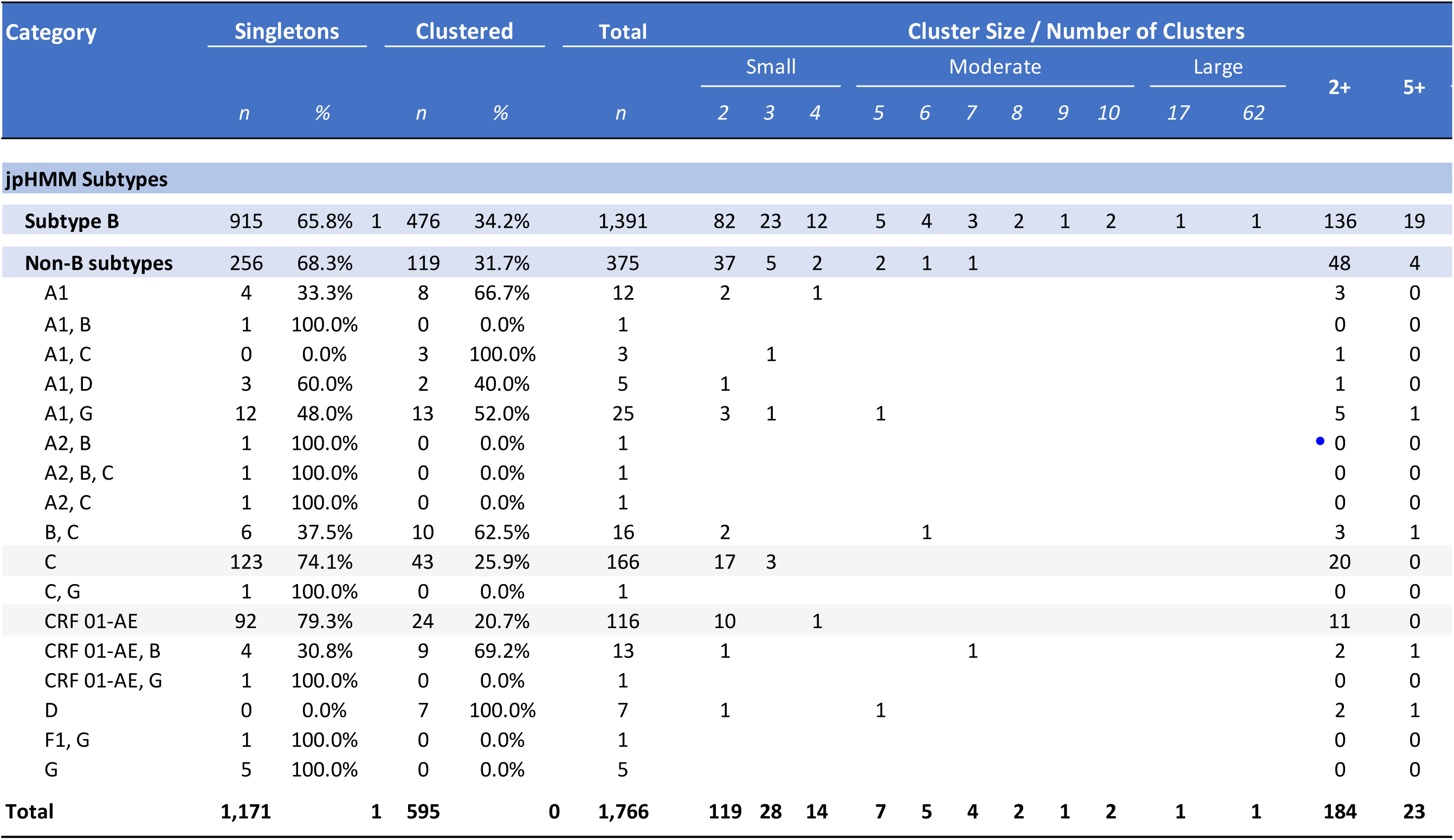
MEGA clusters and their size with patristic distance of ≤1.5% by HIV-1subtype and sequence linkage.

### HyPhy Analysis

The total duration of the analysis of the 1766 study sequences with HyPhy at reference patristic distance ≤2.0% was less than 30 minutes. Figure 2 shows a network diagram of 50 individually outlined clusters including 43 moderate-sized clusters (with 5 to 14 sequences) and seven large clusters (with 15+ sequences) with individual sequences displayed according to HIV-risk exposure. Each node (sequence) in each cluster was customised according to shape (seroconverter status), colour (sex is known), and risk factor. The distance between nodes represents similarity of sequences; shorter lines or a more compact cluster shape with overlapping nodes indicates greater similarity of sequences. Empty (white) circular nodes are sequences without linkage to a notification record. The network diagram in Figure 3 illustrates this more clearly for 7 moderate-sized clusters.

**Figure 2:**
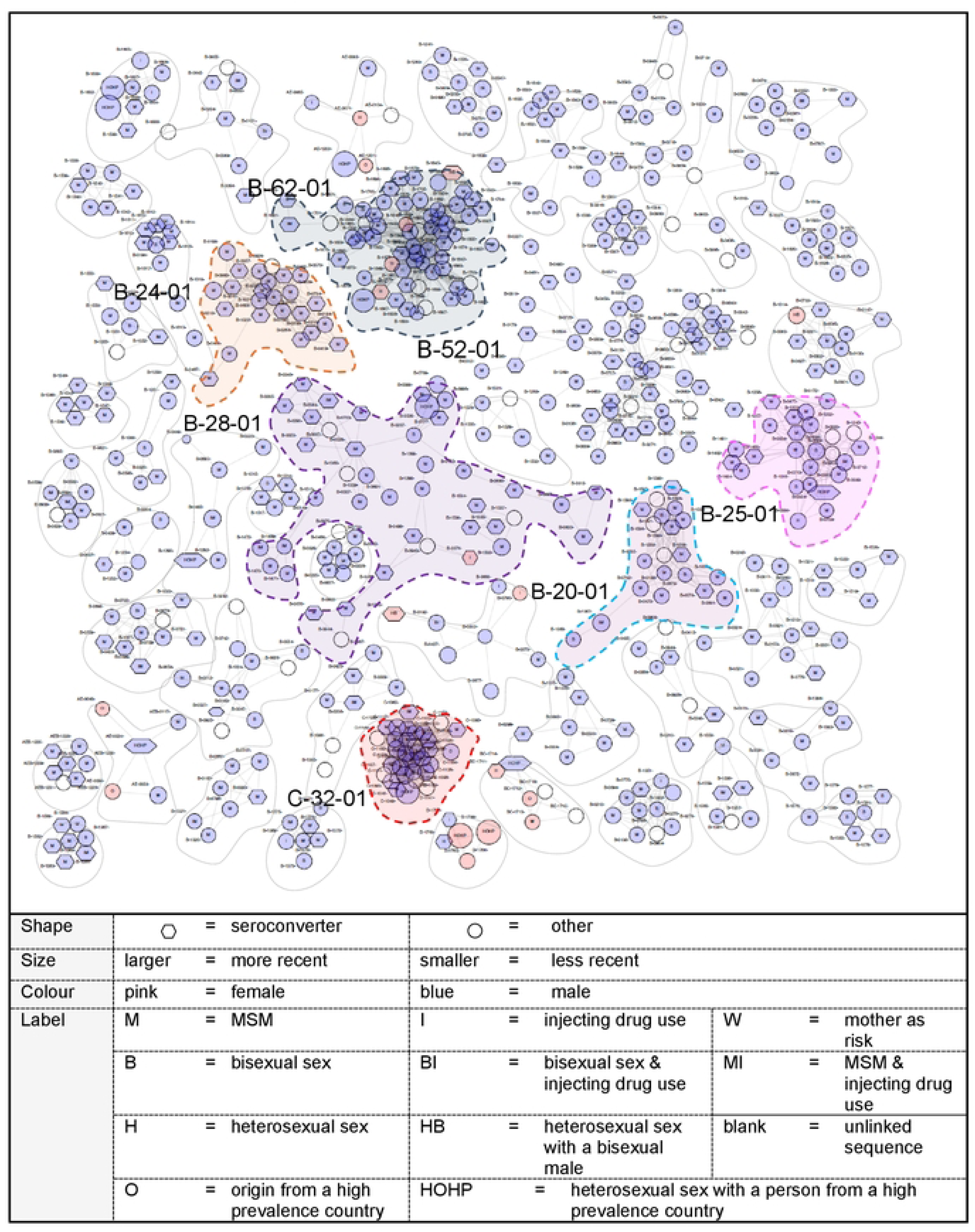
A HyPhy network visualisation of 565 sequences included in 43 moderate-sized clusters (5-14 individuals) and seven large clusters (15 or more individuals). The seven large molecular are highlighted. Separately labelled individual sequences are shown according to HIV risk exposure {detail not visible in this graphic). A lozenge shape indicates an individual who recently seroconverted; grey indicates that sex is known.

**Figure 3:**
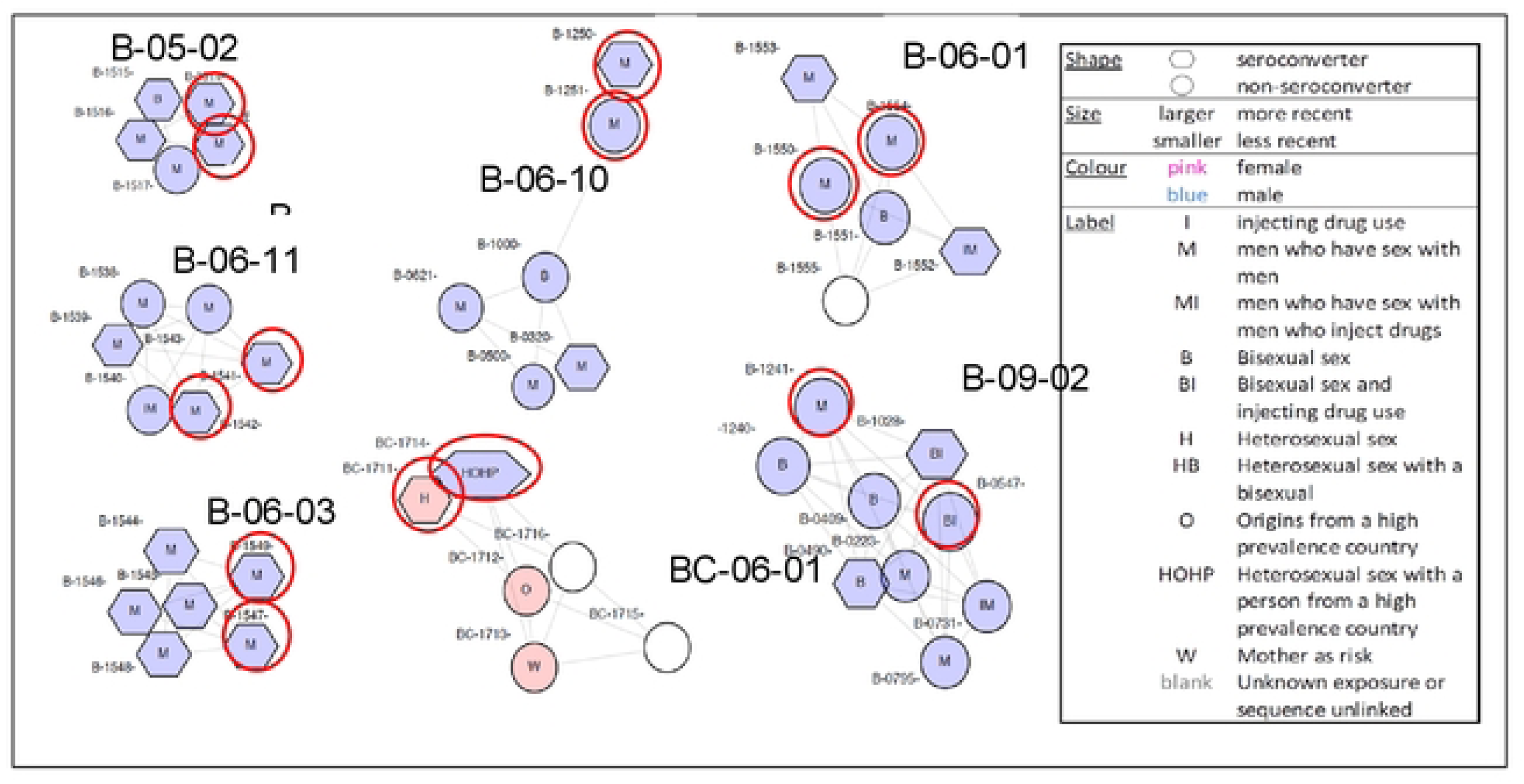
A HyPhy network diagram of seven moderate-sized clusters (5-14 individuals). Pink colour is for female, blue for male. The rings show where at least two new sequences were identified in calendar year 2013. The label denotes HIV risk exposure The distance between nodes represents similarity of sequences - shorter lines indicating greater sequence similarity.

Using ≤2.0% as reference, lowering the patristic distance to ≤1.5% reduced the number of large clusters to six (B-28-01 was no longer detectable), and reduced the size of five clusters (except C-32-01). Reducing it to ≤1.0% reduced the number of large clusters to four and reduced their size. In contrast, raising the patristic distance to ≤2.5% increased the number of large clusters to nine (B-15-01 and B-22-01 became detectable), and increased the size of five clusters (B-20-01, B-24-01, B-25-01, B-52-01, and B-62-01); one cluster was unchanged (C-32-01). Raising it further to ≤3.0% increased the number of large clusters identified from seven to twelve and further increased the size of four clusters (Table 2).

**Table 2:**
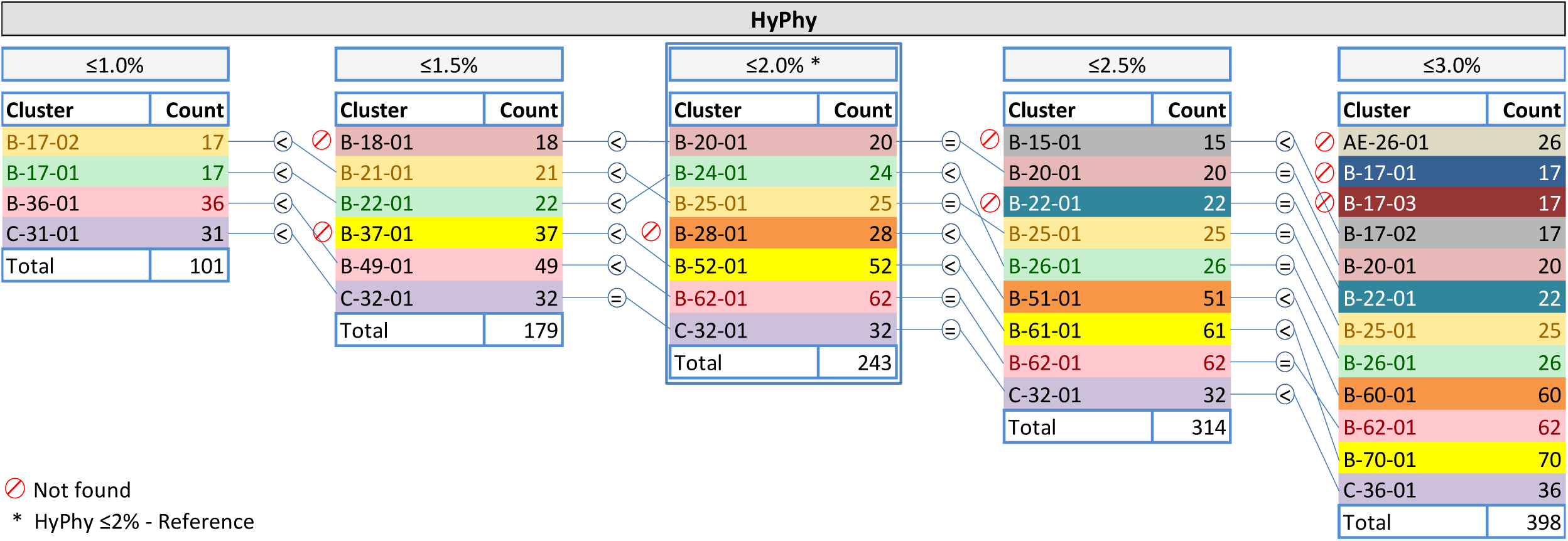
A comparison of the large clusters (15 or more sequences) identified by HyPhy using patristic thresholds between ≤1.0% and ≤3.0%. A patristic distance of ≤2.0% is used as reference. (**3, 20**).

With ≤2.0% patristic distance (reference), the number and size of clusters identified by subtype and sequence linkage and the frequency of singleton sequences, small, moderate and large clusters is depicted in table 3. Overall 1084 (61.4%;1084/1766) study sequences formed 266 clusters of two sequences and 50 larger clusters with five or more sequences with a minority (682; 38.6%) being singletons that were just one individual sequence that did not cluster with any other sequence: we identified 266 pairs, 43 moderate and seven large clusters. There were 253 subtype B clusters including 209 pairs and 44 moderate or large clusters; and 63 non-B subtype clusters including 57 pairs and 6 moderate or large clusters. Of the large clusters one had 53 sequences and one had 62 sequences.

**Table 3:**
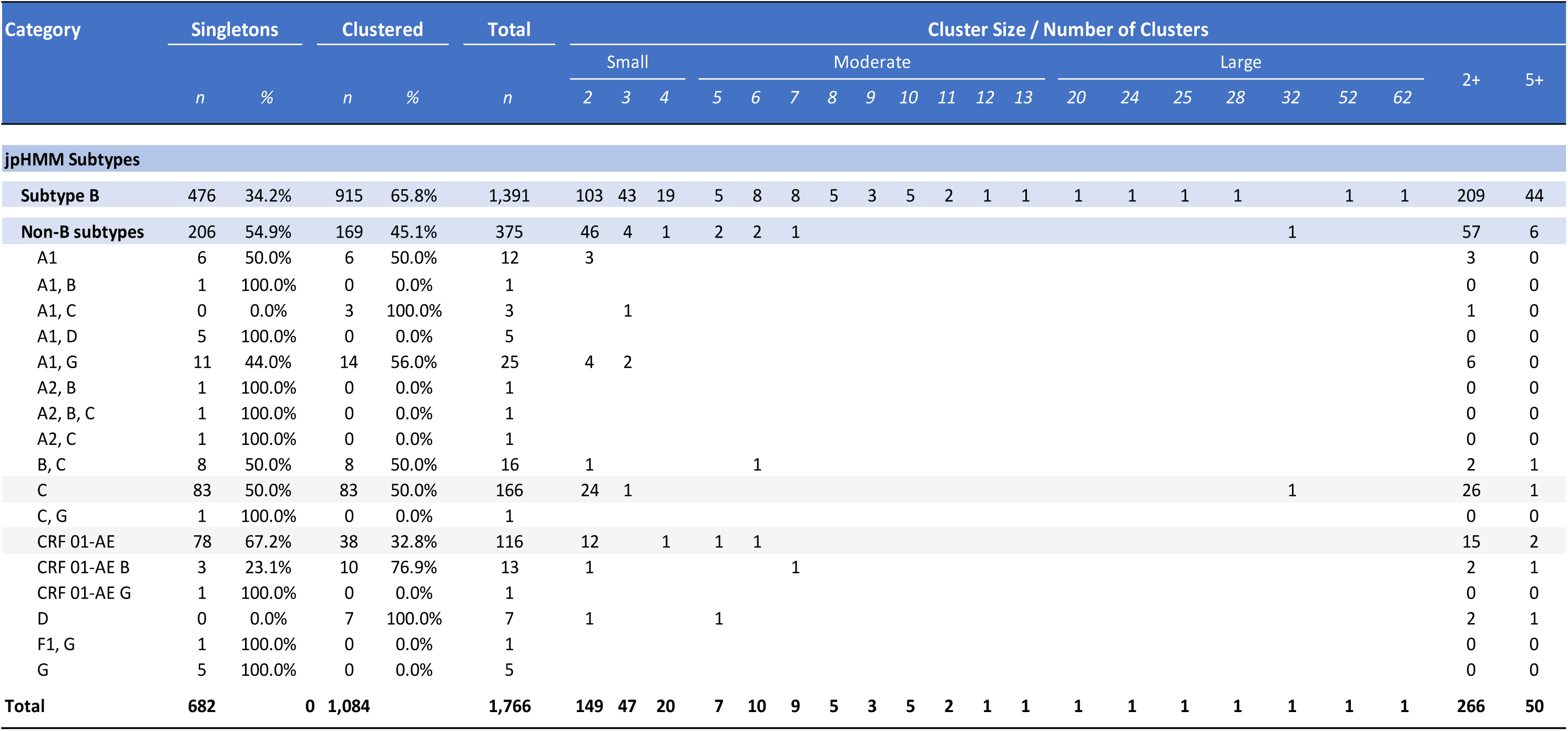
HyPhy clusters and sizes with patristic distance of ≤2.0% (reference) by HIV-1subtype and sequence linkage.

Figure 4a shows a network diagram of one large cluster (B-24-01) with 24 individual sequences of which 23 were linked to a notification record. Within the node, exposure risk factors are shown; all 23 linked sequences were in men who have sex with men. The distance between nodes represents similarity of sequences: shorter lines indicating greater sequence similarity. The cluster first appeared in 2003 and it was still active 11 years later, with three new sequences added in 2013: an average of 2.2 sequences were added to the cluster per year. The timeline for the cluster evolution is shown in Figure 4b. None of this is easily represented in MEGA-derived cluster phylogenetic trees.

**Figure 4a:**
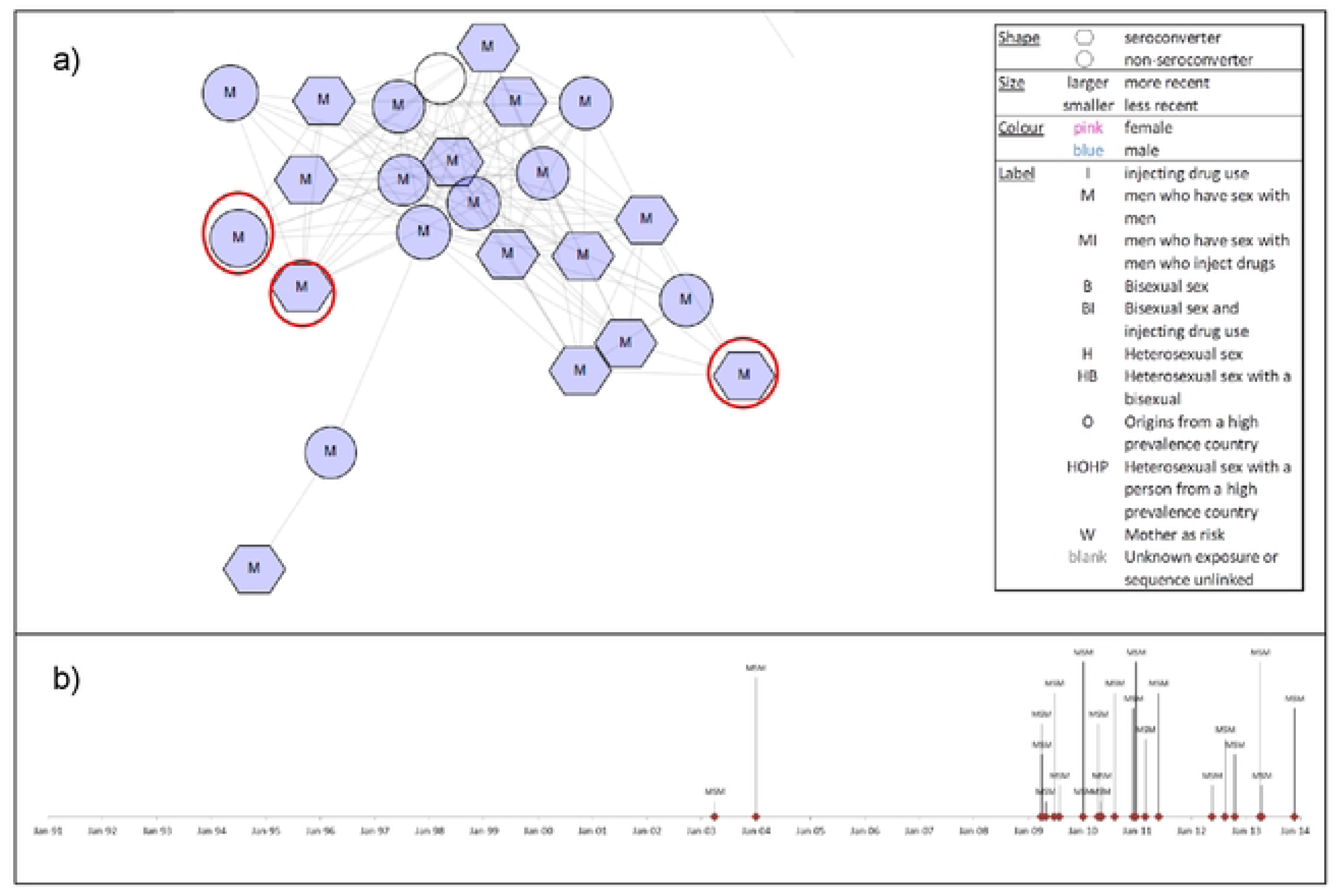
HyPhy network diagram of cluster B-24-01 with 24 unique sequences of which 23 were linked to an individual notification record with patient details as per the box. The distance between nodes represents similarity of sequences - shorter lines indicating greater sequence similarity. The three ringed sequences are the most recent study year 2013. Figure 4b: The timeline for cluster B-24-01 indicating it was first seen in 2003 and still active in 2013, indicating the cluster was of 11 years duration.

### Varying Patristic Distances (Bracketing)

Moderate and large molecular transmission clusters identified using HyPhy ≤ 2.0% patristic distance (reference) were compared with HyPhy ≤ 1.5% patristic distance and MEGA ≤1.5% patristic distance (Table 4). HyPhy ≤ 2.0% was the most efficient and effective, identifying 50 clusters involving 565 unique sequences. Using HyPhy with ≤1.5% patristic distance, 40 clusters with 434 unique sequences were identified. MEGA identified 43 clusters with the smallest number of sequences, just 261.

**Table 4:**
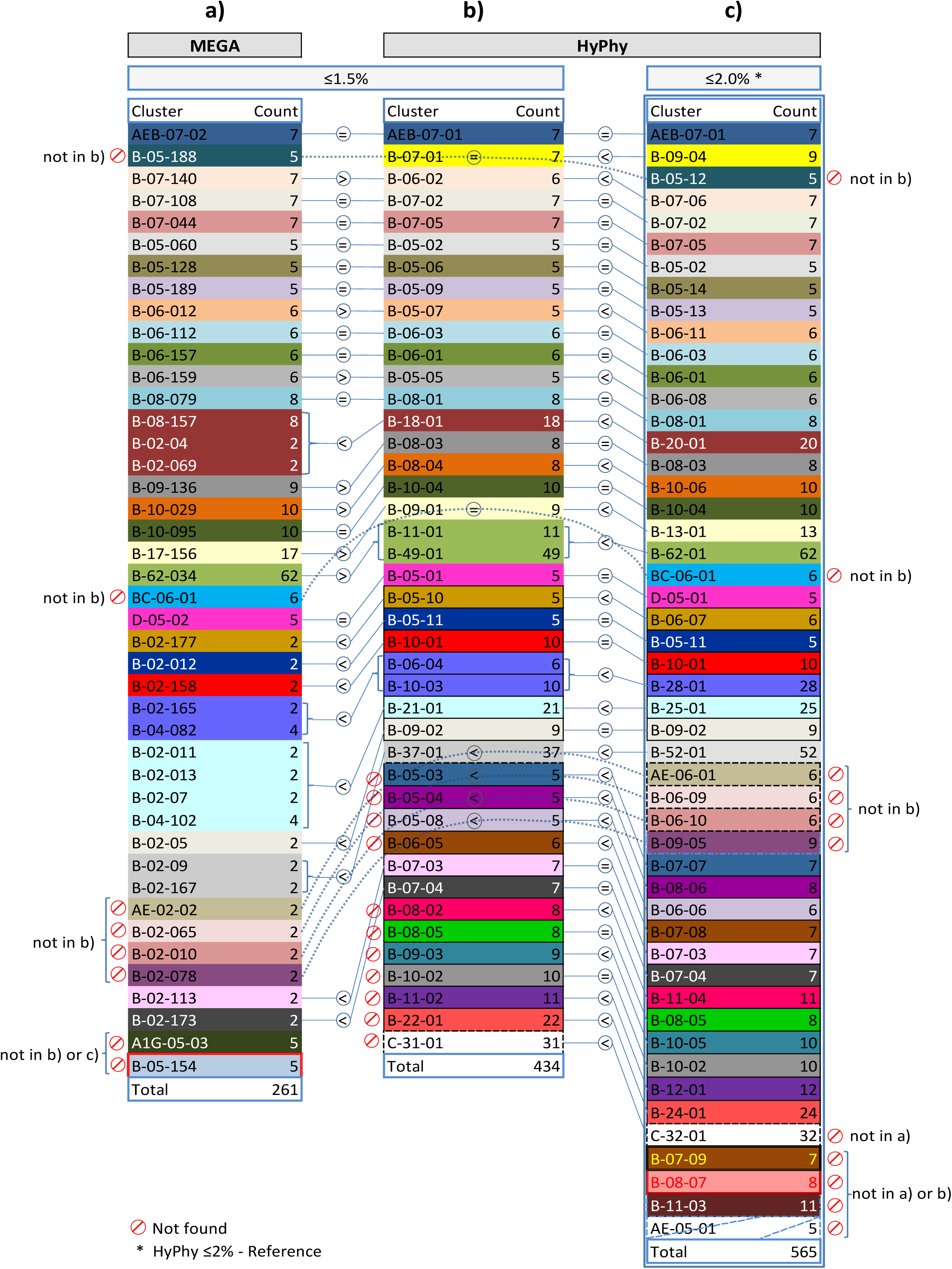
A comparison of moderate (5 to 14 sequences) and large clusters (15 or more sequences) identified using a) MEGA ≤1.5% patristic distance; b) HyPhy ≤1.5% patristic distance and c) HyPhy ≤2.0% patristic distance.

There were 29 clusters common to all three methods including 12 clusters of equal size. Six clusters of equal size were identified by HyPhy ≤ 2.0% and ≤1.5%; and five clusters of equal size were identified by HyPhy ≤1.5% patristic distance and MEGA with ≤1.5% patristic distance. There were five clusters in which the cluster was largest using HyPhy ≤ 2.0% and one cluster which was largest using MEGA ≤1.5% patristic distance. Both HyPhy approaches identified 11 clusters which were not identified by MEGA.

### Comparison of Methods

In objectively comparing the performance of both MPA methods, analysis with HyPhy (30 minutes) was 600 times faster than MEGA (324 hours). With HyPhy ≤ 2.0%, 1084 of the 1766 unique sequences (61.4%) clustered compared to MEGA ≤1.5% where just 595 (33.7%) sequences clustered thus making HyPhy 54% (595/1084) more effective for identifying clustering. Overall, using HyPhy identified 82 more transmission clusters than MEGA (266 versus 184) therefore performing 45% (82/184) more efficiently in our study.

In terms of size, HyPhy found 565 sequences clustered in 50 moderate or large clusters with MEGA finding 261 sequences in 21 moderate and 2 large clusters: of these clusters, 43 were identified by both methods but with MEGA nearly half (20; 47%) of the common clusters were small rather than moderate or large. Additionally, HyPhy identified five clusters not identified by MEGA which in turn identified two clusters, both with five sequences, that were not identified by HyPhy. Three of five clusters just identified by HyPhy were moderate and two were large; both of the clusters just identified by MEGA were moderate. There were five clusters in which the cluster was larger using HyPhy and one cluster which was larger using MEGA. Overall, HyPhy ≤ 2.0% (reference) was also more efficient than MEGA ≤1.5% for establishing cluster size.

A more subjective comparison is how clearly phylogenetic relationships can be represented. The HyPhy network diagrams are more simply able to illustrate molecular transmission clusters than the cluttered circular phylogenetic trees derived from MEGA. Much more information can be included in clear network visualizations of each identified cluster: gender, origin, HIV risk exposure, HIV-1 subtype and size of cluster. A further subjective comparison is the ability to clearly illustrate network dynamics and cluster evolution over time. The timelines and durability of each cluster can simply be shown with the HyPhy molecular network diagrams in a way that is not feasible with the congested MAGA circular phylogenetic trees.

## Discussion

The purpose of our study was to compare two well-established methods for molecular phylogenetic analysis, MEGA and HyPhy, in order to consider which may be better suited for routine use in Queensland for near real-time surveillance of HIV transmission to inform HIV prevention interventions. The study identified 1776 unique sequences of which 1563 were linked to an individual notification record as the baseline or first sequence. Various objective and subjective analyses were conducted to compare the two methods.

Several distinct metrics were used to compare MEGA with ≤1.5% patristic distance with HyPhy ≤2.0% patristic distance, the reference recommended by Poon and colleagues (3,20) and an approach we confirmed worked most efficiently in our setting to best identify molecular transmission clusters. With HyPhy, 1084 of the 1776 unique study sequences (61.4) were found more than once and were considered as clustered whereas just 595 (33.7) sequences clustered with MEGA, making HyPhy 54% (595/1084) more effective for identifying clustering. Overall, HyPhy identified 82 more transmission clusters than MEGA (266 versus 184) and could be regarded in our setting as 45% (82/184) more efficient.

In terms of cluster size, HyPhy was able to identify 565 sequences as belonging to 43 moderate and 7 large clusters whereas MEGA performed less than half as well, identifying just 261 unique sequences in 21 moderate and 2 large clusters. Additionally, HyPhy identified five clusters not identified by MEGA which in turn identified two clusters that were not identified by HyPhy. Once clustering is recognised, HyPhy can also be regarded as being much more efficient for establishing cluster size.

One further objective difference was the time taken to process the data using standard personal computer hardware and software operating environments. Analysis with HyPhy (30 minutes) was more than 600 times faster than MEGA (324 hours). The lengthy analysis time with MEGA will significantly constrain the utility of this method for routine surveillance purposes. In contrast the far shorter duration for using HyPhy to generate molecular transmission clusters strongly indicate that it is far better suited for routine use in HIV surveillance. It will readily permit more frequent analysis of new linked sequence data and for this to be done in near real-time as additional GRT sequences are made available from the reference laboratory.

Subjectively, HyPhy network diagrams and cluster maps can more simply illustrate transmission clusters, include more patient information, show their timelines and are easier to update than the cluttered MEGA circular phylogenetic trees. HIV surveillance is enhanced by updating network cluster maps in near real-time as this allows much easier interpretation of cluster dynamics to reveal transmission hotspots and better target prevention interventions.

No one method is recognised as superior or “gold standard” for HIV-1 MPA to definitively identify individual HIV-1 sequences as part of a transmission cluster or to determine the true size of that cluster. We were only able to compare MEGA and HyPhy, widely used open-source methods for HIV molecular phylogenetic analysis. Both use *pol* sequences that are generated from routinely performed genotypic HIV-1 drug resistance testing as part of standard clinical care.

Other methods have been developed, some of which have used HIV *pol* with *gag* sequences (22, 23) and one study sequenced *gp41* with Cluster Picker (24). The PhyML method has been used by one group in Brazil with both *env* and *pol* sequences (25). MEGA and HyPhy just use *pol* sequences derived from routine clinical practice to identify HIV drug resistance. More resources will be needed to generate *gag, env or gp*41 sequences. With additional funds, further methods could have been included in the study and it is possible that other methods covering sequences other than *pol* would have had comparable or better performance than HyPhy.

Our study used MEGA and HyPhy versions available over a decade ago and it is possible that software developments and updates since then may have improved speed and performance. Additionally there may have been some improvements in the graphical representations generated by the different methods thereby improving their functionality to generate cluster maps to support near real-time HIV surveillance. Any subsequent studies will be able to use software updates. HyPhy ≤2.0% patristic distance will nevertheless likely remain the standard for generating near real-time surveillance data for monitoring HIV transmission networks.

## Conclusion

In our setting, HyPhy objectively performed far better than MEGA in effectively identifying *pol* sequences that clustered and in efficiently determining the number and size of the molecular transmission clusters. Subjectively the network diagrams generated by HyPhy are far simpler ways for illustrating cluster dynamics and their timelines than the circular phylogenetic trees generated by MEGA. The network cluster maps are easier to update as well, enhancing their potential use in near real-time HIV surveillance.

With a considerably shorter overall duration time for analysis, HyPhy has more potential for use in routine, near real-time surveillance to identify transmission hotspots and thus relatively easily translate molecular phenotypic analysis into public health action. By comparison, the duration of the analysis and the effort required to identify the clusters with MEGA would generally rule out the method for routine surveillance.

In conclusion, we are confident that HyPhy is far better suited for Queensland Health to use than MEGA to generate near real-time surveillance data for monitoring HIV transmission and to translate the phylogeny data into action for improved HIV surveillance.

## Data Availability

All relevant data are within the manuscript and/or supporting information files

## Acknowledgements

The study was conducted with the support and under the auspices of the Queensland Professorial Chair of BBVs and STIs.

